# Y disruption, autosomal hypomethylation and poor male lung cancer survival

**DOI:** 10.1101/2020.06.17.20132951

**Authors:** Saffron A.G. Willis-Owen, Clara Domingo Sabugo, Elizabeth Starren, Liming Liang, Maxim B. Freidin, Madeleine Arseneault, Youming Zhang, Shir Kiong Lu, Sanjay Popat, Eric Lim, Andrew G. Nicholson, Yasser Riazalhosseini, Mark Lathrop, William O.C. Cookson, Miriam F. Moffatt

## Abstract

Lung cancer is the most frequent cause of cancer death worldwide^1^. It is male predominant and for reasons that are unknown also associated with significantly worse outcomes in men^2^. Here we compared gene co-expression networks in affected and unaffected pulmonary tissue derived from 126 patients with Stage IA–IV lung cancer. We observed marked degradation of a sex-associated gene co-expression network in tumour tissue. The disturbance was linked to fractional loss of the Y chromosome and was detected in 28% of male tumours in the discovery dataset and 27% of male tumours in a 123 sample replication dataset. Depression of Y chromosome expression was accompanied by extensive autosomal DNA hypomethylation. The male specific H3K4 demethylase, *KDM5D*, was identified as an apex hub within this co-expression network. Male patients exhibiting relative tumour *KDM5D* deficiency had an increased risk of death in the discovery dataset (Hazard Ratio [HR] 3.80, 95% CI 1.40 – 10.3, *P*=0.009) and in an independent sample of 1,100 male lung tumours (HR 1.67, 95% CI 1.4-2.0, *P*=1.2⨯10^−10^). Our findings identify tumour-specific weakening of male-specific expression, in particular deficiency of *KDM5D*, as a common replicable prognostic marker and credible mechanism underlying sex disparity in cancer.

## Main

Consistent sex differences in lifetime risk and survival are recognised across many common cancers^3^. In the UK, in 2010, all-cancer incidence and mortality following age adjustment were 14% and 37% higher in males respectively rising to 46% and 53% when considering lung cancer alone^4^. In line with a convergence of smoking habits, the gap between male and female lung cancer incidence rates is narrowing. Nevertheless, males continue to demonstrate an excess of cases and a relative survival disadvantage. Males with lung cancer have an increased risk of death at 5 years compared with females irrespective of stage, age, period of diagnosis and histologic type^2,5^. The mechanisms responsible for worse outcomes in males have not been established but appear to be independent of cigarette smoking, co-morbidities and treatment type^6^.

An abundance of gene expression changes accompanies lung cancer. The scale and diversity of these changes however, have made it difficult to discern central pathogenic processes and their relationship with prognosis. In the present study we therefore analysed gene expression at a system level, comparing transcriptome organisation between NSCLC (non-small-cell lung cancer) tumours and matched unaffected pulmonary tissues through Weighted Gene Co-expression Network Analysis (WGCNA)^7^. We observed a strongly modular organisation amongst expressed genes, including a combination of shared (pulmonary consensus) and divergent (tumour or normal tissue-specific) co-expression networks. We were then able to relate these co-ordinated gene cliques to inter-individual differences in patient attributes including sex.

Human whole transcriptome data were generated from pulmonary tumours and, with few exceptions, matched unaffected tissue (referred to hereon as ‘normal’) using the Affymetrix HuGene 1.1 ST microarray. Tumour samples and adjacent normal lung tissue were donated from surgical resections, undertaken with curative intent. Following quality control, a total of 18,717 transcripts and 237 samples were available for analysis (Table 1). These samples originate from 126 patients and two major NSCLC histological subtypes; lung adenocarcinoma (LUAD) and lung squamous cell carcinoma (LUSC).

**Table 1:** Dataset demographics. Table 1 displays discovery and replication dataset demographics. Information not available is shown as ---, age is expressed in years and deceased is as of the time of last follow-up. Abbreviations: LUAD (lung adenocarcinoma), LUSC (lung squamous cell carcinoma), T (true), F (false), NS (never smoker), EX (ex-smoker), CS (current smoker), NR (not recorded).

|  |  | Normal | Tumour | Age | Sex | Tumour stage n | Smoking n | Deceased n |
| --- | --- | --- | --- | --- | --- | --- | --- | --- |
| | | n | n | $\mu$(sd) | % Male (n M/F) | IA/IB/II/IIA/IIB/III/IIIA/IIIB/IV (NR) | NS/EX/CS (NR) | |
| Discovery | *LUAD* | 83 | 92 | 68.46 | 49.14% | 54/37/0/22/12/0/42/0/7 | 23/90/58 | 77/95 |
|  |  |  |  | (8.92) | (86/89) | (1) | (4) | (3) |
|  | *LUSC* | 30 | 32 | 69.63 | 64.52% | 22/12/0/9/11/0/8/0/0 | 0/39/23 | 33/29 |
|  |  |  |  | (6.79) | (40/22) | (0) | (0) | (0) |
|  | *Overall* | 113 | 124 | 68.77 | 53.16% | 76/49/0/31/23/0/50/0/7 | 23/129/81 | 110/124 |
|  |  |  |  | (8.42) | (126/111) | (1) | (4) | (3) |
| Replication | *LUAD* | 38 | 41 | 64.38 | 36.71% | 14/20/0/21/7/7/2/0/0 |  |  |
|  |  |  |  | (8.6) | (29/50) | (8) | --- | --- |
|  | *LUSC* | 21 | 23 | 68.2 | 75% | 5/13/2/6/6/9/1/2/0 |  |  |
|  |  |  |  | (7.49) | (33/11) | (0) | --- | --- |
|  | *Overall* | 59 | 64 | 65.75 | 50.41% | 19/33/2/27/13/16/3/2/0 |  |  |
|  |  |  |  | (8.4) | (62/61) | (8) | --- | --- |

## Results

### Network structure

By convention, shared and divergent components of transcriptome organisation are specified through the construction of consensus gene co-expression networks, derived from all samples and common across tissues (‘C’)^8^. Comparisons can then be made against networks derived separately in each tissue, allowing identification of tissue-specific networks.

We report 46 networks that demonstrate similar patterns of co-ordination in tumour (‘T’) and histologically normal (‘N’) lung tissue (containing 35 - 881 transcript clusters [TC]), as well as relative conservancy in their higher-order organisation (D(Preserve^*tumour, normal*^) 0.84). More than a third of transcripts (43.4%, n = 8,129) however were not assigned to any consensus network, indicating relative independence or inconsistent patterns of co-ordination between tumour and histologically normal tissues.

Independent network construction in each tissue class yielded 36 networks in tumour samples (33 - 2,220 TC) and 39 in histologically normal samples (34 - 4,756 TC), with a relatively increased fraction of large networks defined here as containing >1,000 transcripts (C: 0% [n = 0], T: 19.4% [n = 7], N: 7.7% [n = 3]). Consistent with a hypothesis of partial tissue specificity, these single tissue analyses resulted in a markedly smaller proportion of transcripts lacking network assignment (T: 11.8% [n = 2,213], N: 3.0% [n = 567]). Specifically, comparison against consensus networks defined one tumour network and five normal networks lacking a clear consensus counterpart (Extended Data Figures 1 and 2 respectively, Fisher’s exact test - log10(*P*)≥10.0).

### Sex-related tissue specificity

One network specific to histologically normal tissue (Normal: lavenderblush3) featured a highly significant relationship with biological sex (bicor 0.82 *P* = 3.72⨯10^−28^, n Obs = 113, see Extended Data Figure 3). Modest relationships with both FEV1 (bicor 0.31, *P* = 3.60⨯10^−03^, n Obs = 85) and BMI (bicor 0.23, *P* = 2.74⨯10^−02^, n Obs = 91) were also observed but did not retain significance when males and females were examined separately, indicating that these associations were mediated by sex. The transcripts comprising this network were significantly enriched for gonosomal (sex chromosome) inheritance (HP:0010985, *P*_adj_ 1.080⨯10^−08^) followed by histone demethylase activity (GO:0032452, *P*_adj_ 1.059⨯10^−05^). The majority of its 39 members (detailed in Supplementary Table S1) mapped to the sex chromosomes (15 X, 16 Y), and its autosomal members (n = 8) also showed prior evidence of sex-biased expression (e.g. *DDX43, NOX5, NLRP2*)^9,10^. These data indicate a theme of sex bias or specificity in normal pulmonary gene expression, in keeping with the known impact of gonadal sex on pulmonary development and physiology.

Almost 95% of this network’s members (37/39 transcripts) lacked assignment to a consensus network, indicating near-complete divergence in co-expression patterning between tumour and histologically normal tissues. Moreover, over 41% of these transcripts (n = 16), in particular those mapped to the Y chromosome (n = 12, 75%), lacked assignment to a tumour network indicating a specific loss rather than restructuring of co-ordination amongst Y chromosome genes in tumour tissue.

The tumour-specific disturbance in sex-related gene co-expression was visualised through hierarchical clustering (Figure 1a). The disturbance could be detected as a discrete branch characterised by a loss or substantial curtailment of male-specific gene expression, comprising more than a quarter of all male tumour samples (n = 18, 28%) including tumours of both an LUAD and LUSC histology. This branch most prominently featured low expression across a cluster of eight Y-chromosome transcripts, as encoded by seven genes (*DDX3Y, EIF1AY, KDM5D, RPS4Y1, TXLNGY, USP9Y* and *UTY*).

**Figure 1:**
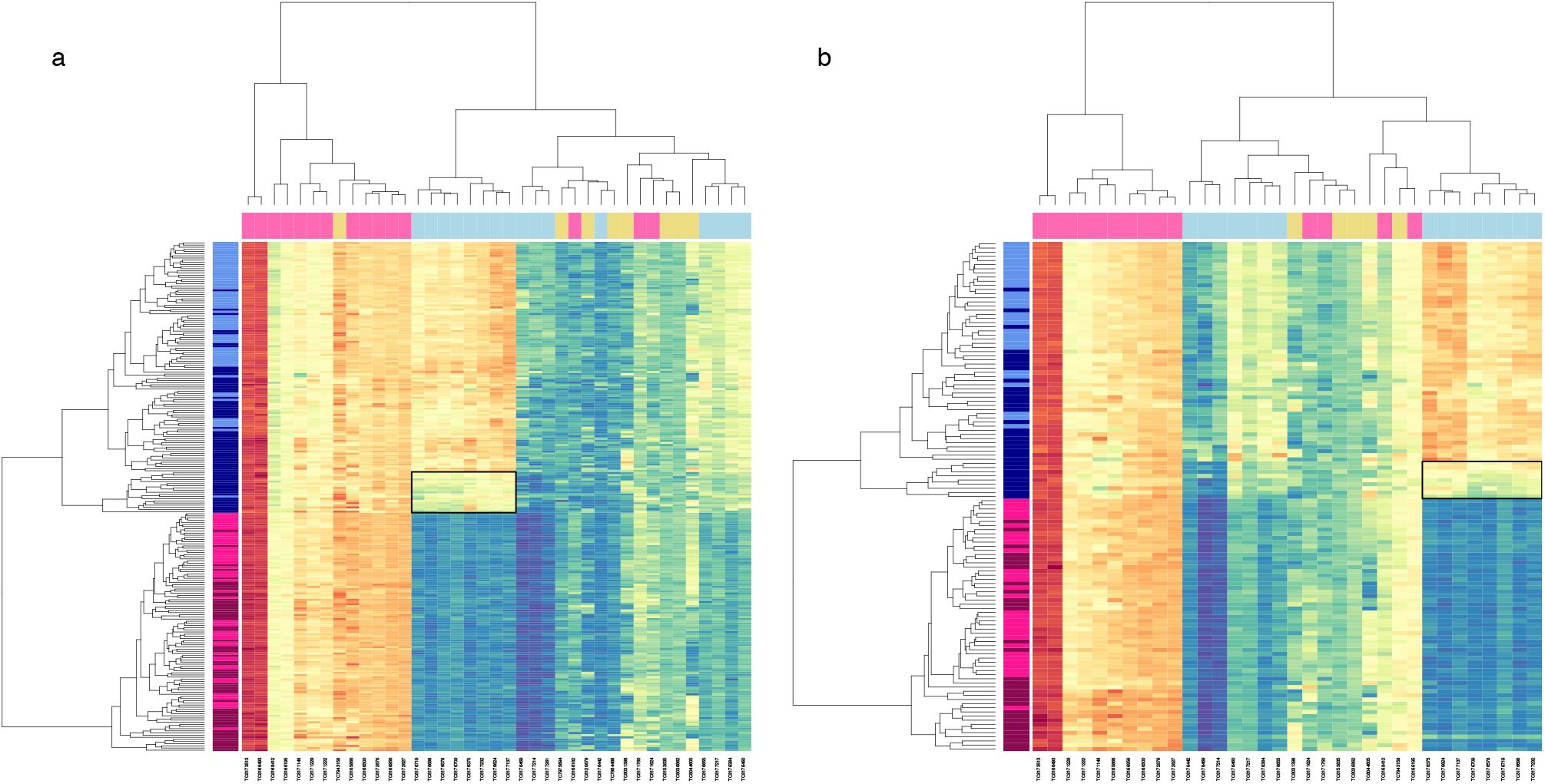
Hierarchical clustering of transcripts assigned to a normal-specific sex associated co-expression network. Hierarchical clustering of transcripts assigned to a normal-specific sex associated co-expression network. Figure 1 displays heat maps with hierarchical clustering of samples (on the Y axis) and transcripts (on the X axis) in discovery (a) and replication (b) datasets, limited to transcripts clusters (TC) assigned to the normal-specific, sex associated, gene co-expression network. Expression is shown on a continuous colour scale from blue (low) to red (high). Sample colour (*y* axis) reflects tissue type (light – histologically normal, dark - tumour) and sex (blue - male, pink - female). Transcript colour (*x* axis) reflects chromosome class (yellow - autosomal, pink – X, blue – Y). Low Y sample / transcript clusters are highlighted by a solid black box.

Data from 34 of the 39 TC comprising the normal-specific sex-associated network were available in an independent sample of 69 lung cancer patients with either LUAD or LUSC, providing 64 tumour and 59 unaffected samples (Table 1). Hierarchical clustering of these 123 samples (Figure 1b) revealed a discrete branch bearing the hallmark of low Y-chromosome expression. The relative depression of Y chromosome expression spanned 8 transcripts, corresponding to 7 Y-chromosome genes (*DDX3Y, EIF1AY, KDM5D, RPS4Y1, TXLNGY, USP9Y* and *UTY*); providing a complete composition match to the discovery dataset. In total the branch contained 9 male tumour samples, representing 27% of all male tumour specimens in the replication dataset.

### Loss of chromosome Y in male tumours

Mosaic loss of the Y chromosome in peripheral blood, concomitant with aging and tobacco smoke exposure^11^, is associated with increased risk for disease and mortality in men^12^ and represents a risk factor for cancer-related mortality^13^. Previous analyses of sex-chromosome aneuploidies have specified six core genes that show obligate Y chromosome dosage sensitivity in their expression^14^. Of these, all 5 available in the discovery dataset (represented on the array and meeting the described filtration criteria) were assigned to the sex-associated network in normal tissue (*TXLNGY* also known as *CYorf15B, DDX3Y, USP9Y, UTY* and *ZFY)* but lacked network assignment in either the tumour-specific or consensus datasets. This indicates a tumour-specific disruption consistent with abnormal Y chromosome dosage.

Somatic loss of Y (LOY) as a mechanism for deficiency of Y chromosome gene expression was queried in the discovery dataset through read depth analysis of whole exome sequencing (WES) and whole genome bisulfite sequencing (WGBS). The subset of male tumour samples exhibiting low Y expression (n WES = 6, WGBS = 17) were compared with matched unaffected tissue from the same patients and with a subset of male tumour samples lacking this feature (n WES = 9, WGBS = 7; Supplementary Tables S3 and S4). Consistent with tumour-specific LOY, normalised read depth was significantly lower in tumours exhibiting low Y-chromosome gene expression as compared with unaffected samples from the same patients (*P*.WES = 0.01082, *P*.WGBS = 2.2⨯10^−16^). This was not the case in male tumours lacking the low Y gene expression signature (*P*.WES = 0.99, *P*.WGBS = 0.97). Correspondingly the percentage loss was significantly greater in males with low Y-expressing tumours than in males lacking this feature (*P*.WES = 0.0266, *P*.WGBS = 0.0460, Extended Data Figures 4a and 4b).

A polymerase chain reaction (PCR)-based chromosome deletion detection assay^15^ was used to corroborate LOY across 20 specific regions of the Y chromosome in 16 patients with low Y expressing tumours (inclusive of the 6 assayed through whole exome sequencing). Relative amplification of these Y-chromosome-specific loci was compared against the expression of genes located in the same physical regions, confirming a positive relationship (*SYPR3* - *KDM5D* r = 0.59, df = 28, *P* = 0.0005; *SY14Y* - *ZFY* r = 0.62, df = 28, *P* = 0.0002). Matched tumour-normal data pairings were available for a total of 15 patients. The ratio between amplification indices in tumour and paired histologically normal samples was indicative of partial somatic deletion in all tumours examined (Figure 2).

**Figure 2:**
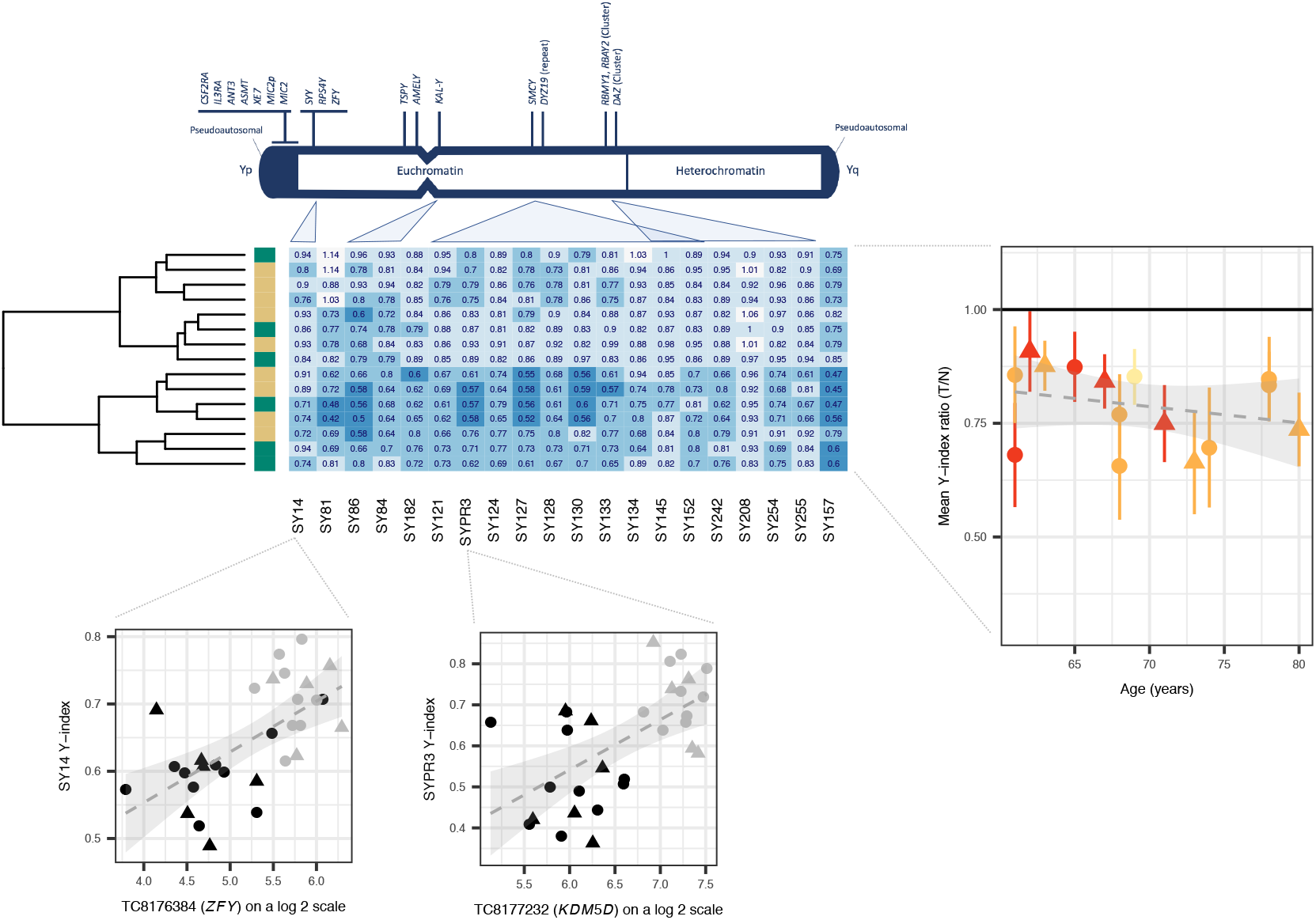
Validation of Loss of Y. Validation of Loss of Y. Figure 2 is headed with a cartoon adapted from the Promega technical manual depicting sites on chromosome Y interrogated through PCR. A heatmap details the ratios between tumour (T) and histologically normal (N) amplification signals on a patient-by-patient basis. Histology is shown as a *y* axis sidebar (LUAD = beige, LUSC = green). The grand mean and standard deviation of these ratios across all sites is plotted against age as expressed in years and coloured by smoking history (never smoker = yellow, ex-smoker = orange, current smoker = red). A hatched linear smooth line is shown with its 95% confidence intervals shaded in grey. The relationship between amplification signal (the Y-index, presented on the *y* axis) and the expression of genes in the same region (presented on the *x* axis) are shown below, with individual points coloured by tissue class (tumour = black, histologically normal = grey) and including a hatched linear smooth line, with 95% confidence intervals shaded in grey. Tumour histology is denoted by point shape (LUAD = circle, LUSC = triangle).

### Autosomal hypomethylation and LOY

Within the sex-associated gene co-expression network, network membership (MM, a metric closely related to intra-network connectivity) was highest for the gene *KDM5D* (MM 0.99, *P* = 6.21⨯10^−94^) (Supplementary Table S1). KDM5D is a male-specific demethylase with established roles in the epigenetic modification of sexually dimorphic histone methylation marks^16,17^. Specifically, KDM5D demethylates trimethylated H3K4 (H3K4me3). This chromatin landmark is generally detected near the start site of transcriptionally active genes^18^ and is anti-correlated with DNA methylation^19^. KDM5D-mediated H3K4 demethylation is required for sex-dependent regulation of gene expression in mouse embryonic fibroblasts^20^.

Consistent with this role, male tumours exhibiting the low Y gene expression phenotype were accompanied by a distinctive autosomal DNA hypomethylation signature (Figure 3). Specifically, median autosomal DNA methylation levels were significantly reduced in these tumours relative to paired unaffected tissues (n 17, *P* = 3.116 ×10^−6^). This relative reduction was not reproduced in male tumours lacking the low Y gene expression feature (n 5, *P*=0.0625) indicating that extensive hypomethylation is a specific characteristic of the low Y pulmonary tumour state and potentially therefore also a latent factor contributing to lung cancer-related the methylation changes reported elsewhere^21^. Autosomal DNA methylation levels were also significantly lower in male tumours exhibiting low Y gene expression as compared with other male tumours lacking this feature (*P*=0.0082). These results demonstrate coincidence between reduced Y chromosome gene expression and widespread autosomal DNA hypomethylation and suggest deficiency of the epigenetic modifier *KDM5D* as a potential mechanism.

**Figure 3:**
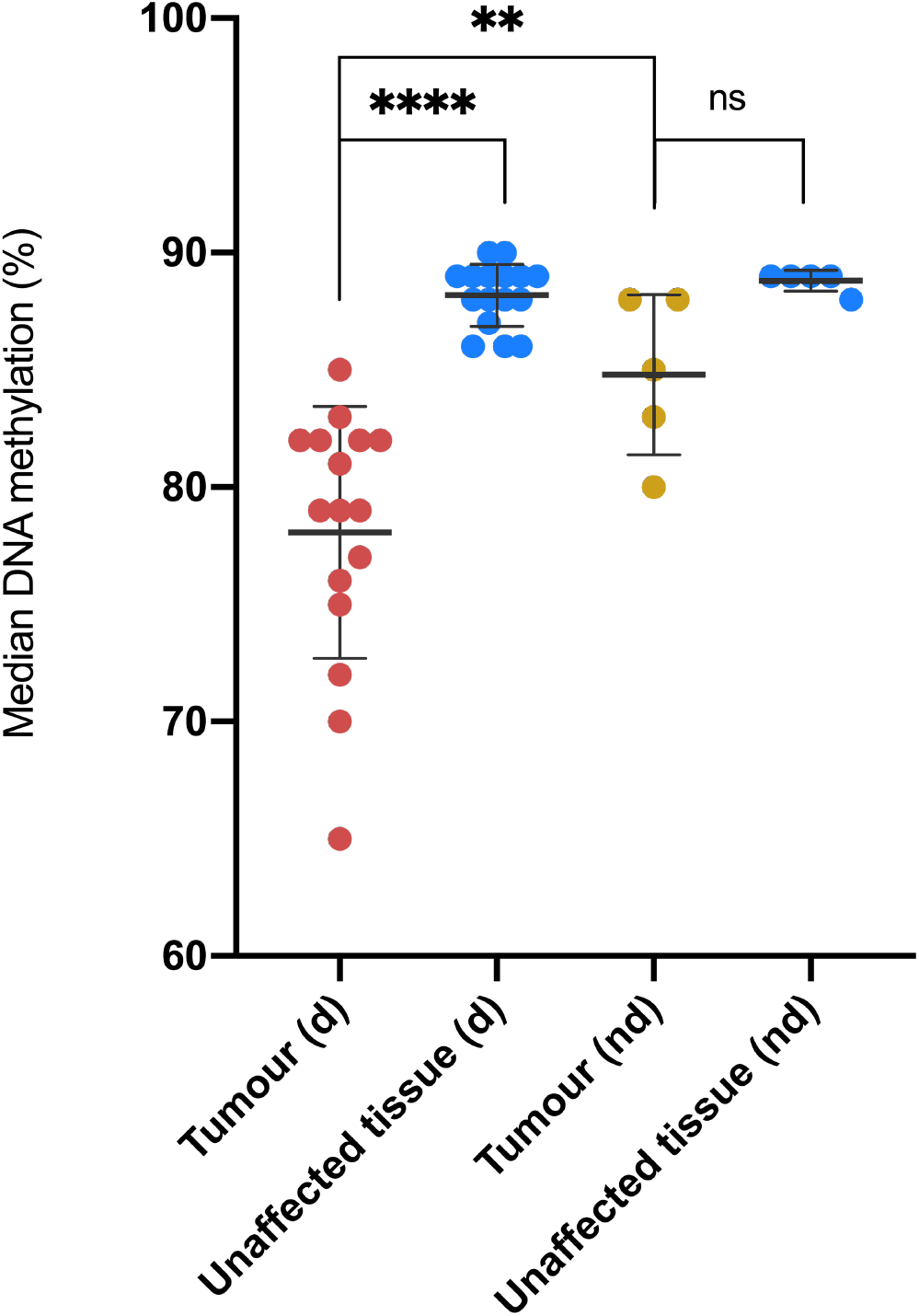
Median DNA methylation percentage per sample. Median DNA methylation percentage per sample. The figure shows median DNA methylation percentage per sample in males with deficient Y chromosome gene expression (d) and males lacking this feature (nd), see Figure 1. Data is shown for both tumour and histologically normal tissue. Normality was assessed with a Shapiro Wilk test. Differences in DNA methylation between paired tumour and histologically normal tissues were assessed using a two-tailed paired t-test (low Y group), and a Wilcoxon test (non-low Y group). A two-tailed unpaired Mann-Whitney test was used to assess differences in DNA methylation between the two tumours groups. Error bars represent standard deviation from the mean. Magnitude of significance is denoted with asterisks (*). Abbreviations: d (deficient chromosome Y gene expression), nd (non-deficient chromosome Y gene expression), ns (non-significant).

Examination of individual regions showing significant differential methylation between low-Y expressing tumours and unaffected paired tissues confirmed cancer-associated changes in DNA methylation strongly biased in favour of hypomethylation. Promoter regions 1Kb upstream of 1,728 genes were hypomethylated in low Y expressing tumours with methylation differences exceeding 20%. These regions showed significant enrichment for multiple motifs relating to the dimeric AP-1 (activating protein 1) transcription factor complex (Supplementary Table 5) which has established roles in malignant transformation and invasion^22^. Nevertheless, hypomethylation was not universal and a total of 473 promoter regions were significantly hypermethylated in low Y expressing tumours. These sites showed significant enrichment for an X-box motif, recognised by RFX transcription factors, and functioning in cellular specialization and terminal differentiation with particular relevance to ciliogenesis^23^.

### Regulation of XY dosage

*KDM5D* has a functional ancestral homolog on the X chromosome, *KDM5C*, which escapes X-inactivation and shows a male-biased pattern of deleterious mutations which associate with male cancer^24^ and multi-locus loss of DNA methylation^25^. We show here that transcript abundance of *KDM5C* is significantly higher in male tumours exhibiting low *KDM5D* expression (≥1.5 SD below the overall male mean) as compared with male tumours lacking this feature (n 65, W = 181, *P* = 0.01). These data indicate active regulation of the dosage balance between these gametologs and suggest that overexpression of *KDM5C* is unable to fully compensate for deficiency of *KDM5D*.

### Prognostic value of tumour *KDM5D*

Down-regulated expression of *KDM5D* has previously been reported in the context of renal cell carcinoma^15^, prostate cancer^26^ and gastric cancer^27^; in at least a proportion of tumours due to somatic loss or segmental deletions of the Y chromosome. Clinically, low *KDM5D* expression is variably associated with a poorer prognosis, more aggressive phenotype and metastasis.

At the time of last follow-up 33 male patients with tumour samples had died. Of these, 8 (24%) had markedly low male tumour *KDM5D* expression (≥1.5 SD below the overall male mean), meaning that almost two thirds (62%) of all males with low tumour *KDM5D* had died as opposed to 49% of males lacking this marker.

We sought to isolate the relationship between *KDM5D* deficiency and prognosis in lung cancer by fitting a multivariate Cox proportional hazards model in the discovery dataset (Figure 4). Following adjustment for baseline prognostic and epidemiological covariates including age, sex, histology, smoking history and tumour stage, markedly low tumour *KDM5D* expression in males was associated with an increased relative hazard of death as compared with females or males with normal range *KDM5D* (HR 3.80, 95% CI 1.40 - 10.3, *P* = 0.009). Significance was retained in an equivalent analysis restricted to males only (HR 4.92, 95% CI 1.464-16.6, *P* = 0.01).

**Figure 4:**
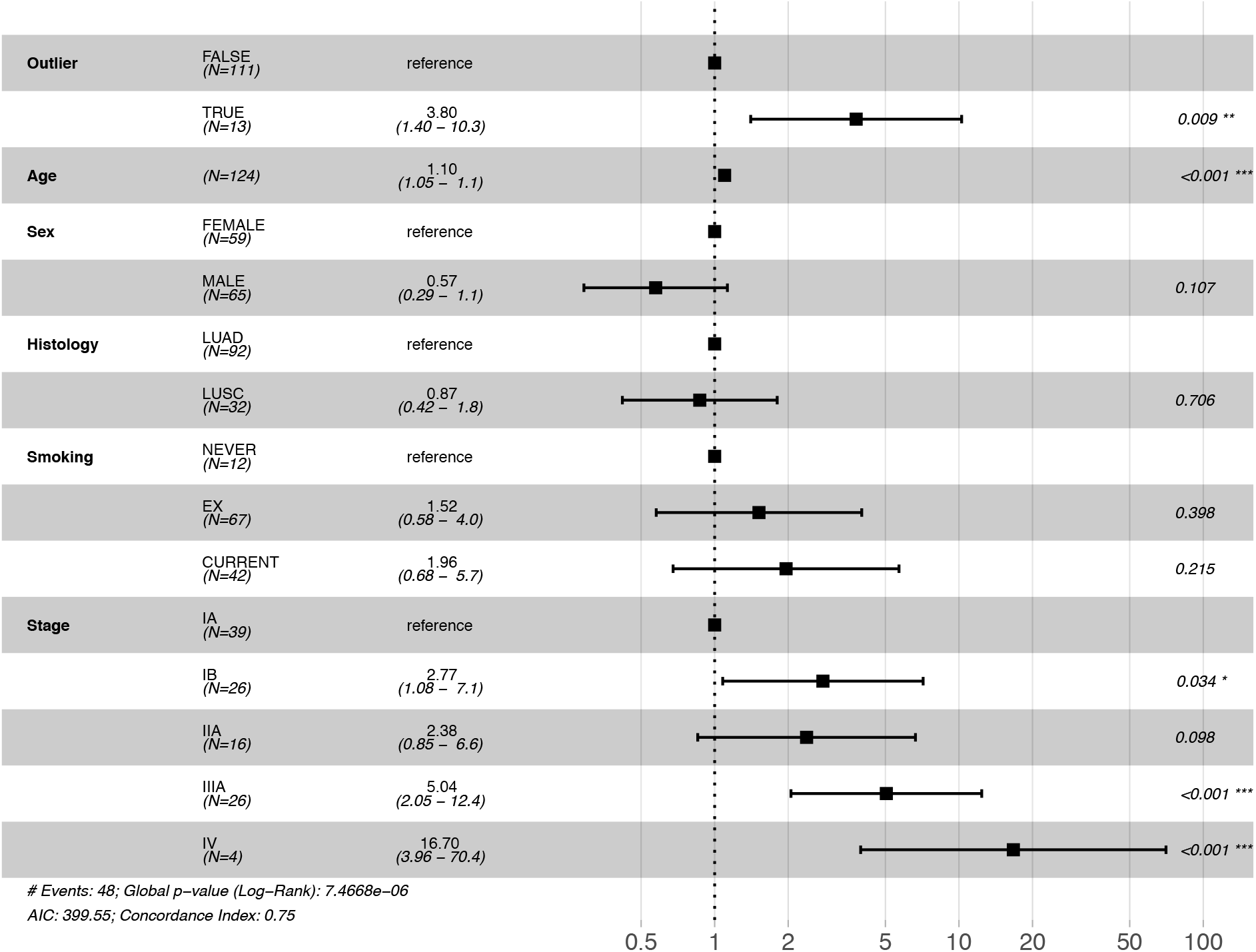
Forest plot for Cox proportional hazards model. Forest plot for Cox proportional hazards model. The figure provides a forest plot reporting the hazard ratio (HR) and the 95% confidence intervals of the HR for each covariate included in the Cox proportional hazards model. The variable *Outlier* specifies male tumour samples showing relative *KDM5D* deficiency (≥1.5 SD below the overall male mean). Abbreviations: LUAD (lung adenocarcinoma), LUSC (lung squamous cell carcinoma), AIC (Akaike information criterion). Magnitude of significance is denoted with asterisks (*).

Notably a model evaluating the wider impact of low Y expression, as indexed by tumour membership of the low Y cluster (shown in Figure 1a), yielded broadly similar (HR 4.19980, 95% CI 1.66 – 10.59, *P =* 0.002) although statistically distinguishable results (*P* = 0.0165), indicating the presence of other prognostically relevant effects amongst the Y cluster genes. In silico validation of the association between tumour *KDM5D* and survival was sought via the online Kaplan-Meier plotter platform (http://kmplot.com/analysis/) accessing 1,100 male tumour samples derived from 11 independent lung cancer mRNA gene chip datasets. Consistent with our observations in the discovery dataset, relatively low tumour *KDM5D* mRNA expression was associated with an unfavourable prognosis in males (HR 1.67, 95% CI 1.4 to 2.0, *P* = 1.2⨯10^−10^, Extended Data Figure 5).

### Wider role in male predominant tumours

Tumour *KDM5D* abundance, as gauged through RNA-seq, was available through the Kaplan-Meier plotter platform across 14 non-sex-specific cancer types totalling 2,423 male patients. Survival analysis incorporating low *KDM5D* as a prognostic indicator yielded nominally significant *P*-values (*P* ≤ 0.05) in seven cancers types and most significantly in head-neck squamous cell carcinoma (HR 1.79 [1.3-2.5], *P* = 0.0003) and liver hepatocellular carcinoma (HR 1.85 [1.16-2.94], *P* = 0.008). Both head and neck and liver hepatocellular carcinoma have raised incidence in males^28,29^, and within head and neck cancer male sex also carries a significant survival disadvantage. We note that the smallest *P*-values were observed in cancers where automatic thresholding placed a cut-off below 20% of the maximum recorded in that tissue (Supplementary Table S2) suggesting a low natural split in the male abundance spectrum in some cancers. Nevertheless, when *KDM5D* abundance was alternatively split at the lowest quartile, significance was retained for both head-neck squamous cell carcinoma (HR 1.75 [1.3-2.5], *P* = 0.0011) and liver hepatocellular carcinoma (HR 1.82 [1.1-2.9], *P* = 0.0099).

### Therapeutic indicators

There is a recognised need for biomarkers capable of predicting systemic therapy sensitivity and response. In prostate cancer (PC) cell lines, low expression of *KDM5D* is already associated with a reduced sensitivity to docetaxel (in the presence of androgen) and cisplatin^30^. Conversely, PC cells deficient for *KDM5D* show an increased sensitivity to ATR inhibitors (ATRi)^26^; compounds that are currently in early phase clinical trials as cancer therapeutics or chemo-sensitizing agents^31^. Specifically, PC cells deficient for *KDM5D* exhibit replicative stress and ATR activation and, following exposure to ATRi, curtailed proliferation and increased apoptosis. This indicates that at least within the context of PC, the combination of *KDM5D* deficiency and ATRi constitute a synthetic lethal interaction^26^.

Whilst extension of these findings to NSCLC or SCLC has not been explored, our findings suggest that LOY-mediated curtailment of Y-chromosome gene expression, particularly deficiency of the demethylase *KDM5D*, may identify a male patient group with distinct progression, mortality and synthetic lethality profiles.

## Data Availability

Gene expression data have been deposited in NCBI's Gene Expression Omnibus and will be accessible through GEO SuperSeries accession number GSE151103 (https://www.ncbi.nlm.nih.gov/geo/query/acc.cgi?acc=GSE151103); comprising SubSeries GSE151101 (discovery) and GSE151102 (replication) upon acceptance for peer-reviewed publication. Sequence data are available upon request.

## Methods

### Study cohorts

Tumour samples and adjacent normal lung tissue were donated from surgical resections undertaken with curative intent at the Royal Brompton Hospital between 2009 and 2011 as part of a wider study. Written informed consent for research on biobanked tissue was obtained, with any patients lacking such consent entirely excluded from the study. The study methodologies followed the standards set by the Declaration of Helsinki and were conducted under approval by the Royal Brompton and Harefield Research Ethics Committee (RBH) NIHR BRU Advanced Lung Disease Biobank (NRES reference 10/H0504/9) and Brompton and Harefield NHS Trust Diagnostic Tissue Bank (NRES reference 10/H0504/29) [Discovery], and the Royal Brompton and Harefield Ethics Committee (REC reference number LREC 02-261) [Replication]. Within two hours of resection tissue samples destined for transcriptomics were stored in RNAlater (Qiagen, Crawley, UK) whilst tissue samples for genomic DNA were snap-frozen and archived at −80 °C. Histology was determined through review of pathology reports and examination of haematoxylin and eosin (H&E) stained sections (A. Nicholson).

### Gene expression data pre-processing

#### I. Discovery data set

Gene expression data from the Affymetrix HuGene 1.1 ST array were available for a total of 309 samples. Of these, 6 samples from patients with tumour types individually represented by only a single patient, or lacking appropriate consent for external processing were removed. Quality of the remaining expression data was assessed through arrayQualityMetrics (3.30.0) and the RLE (Relative Log Expression) and NUSE (Normalised Unscaled Standard Errors) metrics calculated within the Bioconductor package Oligo (1.38.0). These metrics highlighted 7 samples (2.3% of the input dataset) as potentially problematic and these were removed. Raw expression data for the remaining 296 samples were RMA-treated using Oligo (1.38.0) and filtered. Specifically, transcript cluster intensity was required to exceed the data set median in 1 or more sample (genefilter 1.56.0), and be designated within the Affymetrix annotation (netaffx build 36) with a cross-hybridisation potential of 1 (unique), a non-missing mRNA assignment and as part of the main design probe set category. Together these filters yielded 18,717 transcript clusters (TC). Gene annotations were collated from the netaffx build 36, and the Bioconductor package hugene11sttranscriptcluster.db (8.5.0) as assembled from public repositories. Samples derived from patients with a LUAD or LUSC histology were selectively retained for analysis (Table 1).

#### II. Replication data set

Gene expression data from the Affymetrix HuGene 1.1 ST array were available for a total of 123 samples from 69 patients with either a LUAD or LUSC histology. Quality control and data pre-processing were carried out as described for the discovery dataset, yielding a final data dimension of 123 samples and 17,264 TC.

### Gene co-expression network analysis

A consensus network analysis of tumour and normal lung expression data was performed using step-by-step unsigned WGCNA (1.51)^32^, employing a soft-thresholding power of 5 (Extended Data Figure 6) and scaling topological overlap matrices (TOM) for purposes of comparability (scaling parameter 0.95).

Adaptive branch pruning was performed using dynamicTreeCut (1.63-1), applying a minimum cluster size of 30, a maximum joining height of 0.995 and a deep split parameter of 2 (specifying the sensitivity to cluster splitting). Modules classified as too close in terms of the correlation of their module eigengenes were merged (maximum dissimilarity that qualifies modules for merging 0.25). Consensus modules were related to phenotypic traits through two-sided biweight mid-correlation (robustY=FALSE, maxPOutliers=0.05) as per recommended best practice for settings that include binary or ordinal variables, and compared with modules identified in tumour or unaffected tissue alone as calculated using equivalent computational parameters. Pathway enrichment analysis was implemented in g:profiler (https://biit.cs.ut.ee/gprofiler/)^33^ based on unique Entrez ID annotations (as determined through hugene11sttranscriptcluster.db 8.5.0) and incorporating the tailor-made g:SCS algorithm for multiple testing correction.

### Sequencing

Whole Exome Sequencing (WES) and Whole Genome Bisulfite Sequencing (WGBS) were performed at the McGill Genome Centre, Montreal, Canada. Research samples consisted of genomic DNA extracted from surgically resected, fresh-frozen human lung tumour specimens and normal paired tissue. WES sequencing libraries were prepared with the SureSelect^XT^ Target Enrichment System (Agilent SureSelect Human All Exon V4) and sequenced with Paired-End Illumina HiSeq2000 Sequencing. Non-directional Whole Genome Bisulfite Sequencing (WGBS-Seq) libraries were constructed and sequenced with paired-end Illumina HiSeq X Next Generation Sequencing. Both WES and WGBS were performed according to standard protocols.

### Read depth analysis

Sequencing read coverage was analysed for a total of 21 samples (15 patients, described in Supplementary Table S3) through the analysis of WES data (McGill University Innovation Centre, Montreal) available as part of a wider study. Sequence read coverage was obtained for all chromosome Y genes using the BEDtools coverage tool and normalised both by gene length and sample sequencing depth. Percentage of loss of chromosome Y was then calculated considering only the captured regions. Normality of the data was examined through Shapiro-Wilk normality tests. Paired and un-paired t-tests were performed as appropriate, to examine between-group differences, and these were plotted using GraphPad Prism (8.3.1).

### Differential Methylation

Whole-Genome Bisulphite Sequencing (WGBS) was used to assess DNA methylation levels in tumour and matched unaffected samples in the low Y expression group of males and in additional males lacking this feature as a control group. Non-directional WGBS libraries were constructed and sequenced with paired-end Illumina HiSeq X Next Generation Sequencing at the McGill University Innovation Centre in Montreal. Analysis of WGBS-Seq data was performed with GenPipes^34^. The standard GenPipe for methylation analysis Methyl-Seq is adapted from the Bismark pipeline. Alignment was performed with bismark (0.18.1) and bowtie2 (2.3.1) according to bismark user guide manual with default options. SAM files thus obtained per sample were sorted by chromosomic location with GATK (Genome Analysis Tool Kit) (3.7) and read alignments deemed to be PCR duplicates were removed with Picard (2.9.0). Bismark methylation extractor was used to extract methylation in CpG context. Methylkit R package (1.12.0) was used to obtain median methylation per sample and clustering based on methylation profiles.

Calling of Differentially Methylated Regions (DMRs) was performed with Dispersion Shrinkage for Sequencing data with single replicates (DSS-single)^35^ implemented in the DSS Bioconductor R package (2.34.0), which takes into account spatial correlation, read depth and biological variation between groups. DMRs were called using the criterion absolute methylation differences >20% and *P* <0.001.

Coordinates 1Kb upstream hg19 Ensembl genes were downloaded from UCSC Table Browser to obtain promoter genomic regions. Proximity of DMRs to promoter regions was analysed with Bedtools’ IntersectbED^36^. Then, enriched TF binding motifs in the genomic regions of promoters employed the motif enrichment algorithm in the HOMER tool^37^. CpG normalization and use of the repeat-masked sequence were the options given for finding enriched motif in the genomic regions given.

### PCR-based detection of LOY

The Y Chromosome Deletion Detection System assay, Version 2 (Promega, WI, USA) was performed in a total of 16 patients from the low Y tumour group (31 samples, 15 complete tumour-normal tissue pairs), across 20 regions of the Y chromosome as per the manufacturer’s instructions and as detailed elsewhere^15^. Briefly, the intensity value for each Y-linked amplicon was normalized to the intensity value of corresponding (non-Y) control amplicon obtained from the same sample. The average of these values across 3 replicates, the Y-index, was used to calculate a patient-specific tumour:normal ratio. Corresponding expression data were available for all but one of these samples.

### Survival analysis

Survival curves and a multivariate Cox proportional hazards model were fitted using the R package Survival (2.44-1.1). Survival curves and forest plots were drawn using survminer (0.4.3). Model comparison was achieved through an implementation of the likelihood-ratio test for Cox regression models as proposed by Fine^38^ (nonnestcox 0.0.0.9000).

### In silico validation of tumour *KDM5D* as a prognostic marker

The prognostic value of tumour *KDM5D* in male cancer was assessed via the Kaplan-Meier plotter (http://kmplot.com/analysis/); an online platform providing access to overall survival data in combination with gene chip or RNA-seq transcriptional data^39,40^.

#### I. Lung cancer

Arrays designated as biased through the Kaplan-Meier plotter quality control pipeline were excluded. Overall survival was available in 1,100 male patients with lung cancer split across 11 independent cohorts (CaArray, GSE14814, GSE19188, GSE29013, GSE30219, GSE31210, GSE31908, GSE37745, GSE4573, GSE50081 and TCGA). *KDM5D* was accessed through the Affymetrix ID 206700_s_at (range 3 - 3581) with automatic thresholding (applied cut-off 515, 14.38% of maximal).

#### II. Pan-cancer

The wider prognostic value of tumour *KDM5D* in male cancer outside of the lung was explored via the Kaplan-Meier plotter utilising RNA-seq data available across a total of 2,423 male patients and 14 cancer types, excluding sex-specific cancers and cancers individually represented by ≤20 samples. These included bladder carcinoma (n = 298), esophageal adenocarcinoma (n = 69), esophageal squamous cell carcinoma (n = 69), head-neck squamous cell carcinoma (n = 366), kidney renal clear cell carcinoma (n = 344), kidney renal papillary cell carcinoma (n = 211), liver hepatocellular carcinoma (n = 249), pancreatic ductal adenocarcinoma (n = 97), pheochromocytoma and paraganglioma (n = 77), rectum adenocarcinoma (n = 90), sarcoma (n = 118), stomach adenocarcinoma (n = 238), thymoma (n = 62) and thyroid carcinoma (n = 135). Automatic thresholding was applied.

## Code availability

Network analysis code utilised in this manuscript follows the publicly available WGCNA consensus pipeline^32^. Scripts used for sequence analysis are available upon request.

## Acknowledgements

We thank the patients for participating to our study. This study was supported by the Asmarley Trust and by the Wellcome Trust. Sample collection for this study was supported by the NIHR Respiratory Disease Biomedical Research Unit at the Royal Brompton and Harefield NHS Foundation Trust. Y.R. is a research scholar of the Fonds de recherche du Québec – Santé (FRQS). The views expressed in this publication are those of the authors and not necessarily those of the NHS, the National Institute for Health Research or the Department of Health. The funding sources had no involvement in study design, collection, analysis or interpretation of the data, or in submission for publication.

## Author Contributions

WC and MM designed and conceptualised the study. ES performed patient recruitment and sample preparation with input from EL and AN and supervision from MM and WC. ES, SL, YZ and MF carried out the microarray processing. CD and ML performed the exome sequencing and read depth analysis. MA and YR performed PCR-based LOY validation experiments and associated data analysis. SW-O performed the transcriptomic data analysis with input from LL. SW-O wrote the manuscript with input from WC, MM, SP and ML.

## Additional Information

### Supplementary Information

Supplementary Information is available for this paper.

